# Diagnostic performance of Xpert MTB/RIF Ultra assay for tuberculosis in stool specimens among adult presumptive TB patients in a generalized HIV epidemic setting

**DOI:** 10.64898/2026.08.20.26360877

**Authors:** Htet Ko Ko Aung, Sein Sein Thi, Wanitda Watthanaworawit, Aung Pyae Phyo, Francois Nosten

## Abstract

**BACKGROUND:** Diagnosis of Tuberculosis (TB) from stool specimen using the Xpert MTB/RIF Ultra assay (Xpert-Ultra assay) is important to confirm diagnosis for presumptive TB patients who are unable to produce sputum. We evaluated diagnostic performance of the Xpert-Ultra assay in stool specimen among adult migrant population living in generalized HIV epidemic situation.

**METHODS:** A prospective, cross-sectional study was conducted at outpatient and inpatient departments of the Shoklo Malaria Research Unit (SMRU) clinics and Mae Tao Clinic (MTC) located in Thailand-Myanmar border area. Presumptive TB patients of any age who were registered between November 14, 2022, and May 23, 2023, were eligible for inclusion based on reported signs and symptoms and/or radiological findings. Using liquid MTB culture in sputum as reference standard, evaluation of diagnostic performance of the Xpert-Ultra assay in stool was performed, and it was also compared with performance of smear microscopy and Xpert-Ultra assay in sputum specimen.

**RESULTS:** Total 113 participants were included in the analysis; 9 (7.96 %) had human immunodeficiency virus (HIV) infection, and 31 (27.43%) had confirmed TB on culture results. Among these culture-confirmed TB cases, the sensitivity of Xpert-Ultra assay in stool specimen was 90.32 % (95% confidence interval [CI], 74.25% to 97.96%). Although the absolute difference in sensitivity of Xpert-Ultra assay in stool was 3.23 % lower than sputum (95% CI: -9.46 % to 3.00 %), there was no statistically significant difference between the two sample types. The specificity of Xpert-Ultra assay in stool specimen was 98.78% (95% CI, 93.39% to 99.97%) against culture-negative TB cases, giving an absolute difference of 1.22 % (95% CI, -1.16% to 3.59%) compared to sputum Xpert-Ultra assay. This method demonstrated that diagnostic performance was consistent with World Health Organization (WHO) target product profiles on low-complexity assays for detecting *Mycobacterium tuberculosis* (MTB).

**CONCLUSIONS:** The Xpert-Ultra assay in stool specimen can be considered as a potential, alternative method in diagnosis of presumptive pulmonary TB in adults when respiratory sample is difficult to collect.

## Introduction

TB remains a significant global health issue and stands as the top leading cause of death among infectious disease worldwide. In 2023, an estimated 10.8 million people were affected by TB disease, and 1.2 million succumbed to death despite curable and preventable [1]. The space of innovation in TB diagnostics has become faster than ever in the 21^st^ century and the WHO endorsed various kinds of rapid molecular diagnostics including near point-of-care tests that are feasible to use in peripheral health facilities without needing sophisticated laboratory infrastructure or highly skilled professional [2]. Yet, one out of four TB patients are still missing to get diagnosed and treated [1] due to a large gap in access to diagnosis and treatment.

Diagnosis of pulmonary TB (PTB) relies mainly on sputum specimens which are often difficult to obtain from children, elderly, people living with HIV, and very sick patients. Additionally, the sensitivity for the sample obtained from patients with paucibacillary TB including children [3], people living with HIV (PLHIV) with low CD4 count [4] and elderlies [5] is low. To improve sensitivity in such population, invasive procedures including nasogastric aspiration, sputum induction, bronchoalveolar lavage are required to obtain specimens. These procedures require skilled personnel and equipment that can only be available in higher-level health facilities, creating inequitable access, diagnostic delays and posing infection risks to healthcare workers through respiratory particle generation during the procedures [6–8]. Therefore, alternative non-invasive, accessible, and safe specimen types are needed for diagnosis of PTB [7–9]. Stool is an alternative specimen for TB diagnosis, because MTB can be swallowed with the sputum and detected in stool [8]. WHO has endorsed stool-based TB diagnosis in children in 2021 [2]. In 2025, WHO recommended concurrent testing of respiratory and stool sample for diagnosis of TB in children due to incremental diagnostic accuracy [10].

However, the studies on the stool-based TB diagnosis relative to sputum samples in adults exhibited varying sensitivity of 21.4% to 63.9% by culture, 12.1% to 53.9% by smear microscopy, and 69.7% to 100% by PCR assays including Xpert-Ultra assay. The specificity ranged from 61.5% to 100% in all diagnostic tests [11]. Because of the inconsistent results of diagnostic performance parameters, more studies and data are required to contribute to meta-analysis of larger data set to determine the accuracy of stool sample use for diagnosis of PTB. Moreover, there is no published stool-based TB diagnosis study conducted for the population in living in area with high TB incidence and generalized HIV epidemic situation, particularly among migrants and border population who are more vulnerable to TB than general population [12]. Therefore, we conducted a study to determine the diagnostic performance of stool sample for detecting *Mycobacterium tuberculosis* deoxyribonucleic acid (MTB-DNA) in relative to respiratory sample for the diagnosis of PTB among migrants and border population with presumptive TB. Our hypothesis was that MTB-DNA would be identified in stool specimen using the Xpert-Ultra assay and smear microscopy.

## Material and methods

### Study design and setting

This cross-sectional observational study was conducted among presumptive PTB patients at SMRU clinics and MTC located in the Thailand-Myanmar border areas, Tak province of Thailand between November 14, 2022 and May 23, 2023. The burden of TB among migrant population in this province was higher than that of local Thai population [12] and generalized HIV epidemic situation is found among migrant population [13]. Moreover, the migrant population in the border areas has limited access to health services due to their legal status and language barrier, increasing their vulnerability to communicable diseases. The SMRU TB program has provided diagnostics and treatment services to migrant and local ethnic population on the border areas of Tak province since 2010. MTC, SMRU-partnered clinic, is a primary health care centre serving migrant and local ethnic populations along the border [13]. Under the training and supervision of SMRU TB technical team, MTC personnel perform TB screening.

### Study population

Using WHO recommended TB screening tools including both symptoms and chest radiography, participants were systematically screened for TB disease [14]. Individuals of any age presenting with self-reporting presumptive TB symptoms more than 2 weeks duration, including fever, cough, haemoptysis, night sweat, weight loss or failure to gain weight at the out-patient and in-patient departments of study sites were assessed, and chest X-ray (CXR) was taken. Participants who had either presumptive TB symptoms and/or radiographic abnormalities suggestive of TB were eligible for enrolment. Any participants who are unable to communicate or refuse to participate were continued in routine medical services. All participants or guardians were requested to provide a written informed consent before enrolment. All participants were consecutively enrolled at the study sites until required sample size was fulfilled.

### Study procedures

Following enrolment process, demographic information, clinical symptoms, HIV, previous TB history and findings of physical examination were documented by trained study staff with standardized case record form designed for routine programmatic data collection. Once history taking and physical examination were completed, CXR was undertaken at the SMRU TB centers or a designated district hospital on the same day. Interpretation of CXR findings were carried out by an experienced medical doctor for evaluation of abnormal radiological lesions consistent with PTB. Sputum or nasogastric aspirate (NGA) for children who could not produce sputum and stool samples were collected for microbiological analysis. The sputum samples were tested with Xpert-Ultra assay (Cepheid, Sunnyvale, United States of America [USA]), smear microscopy on respiratory tract specimen (RTS) (sputum or NGA) with Ziehl–Neelsen staining and liquid culture on the BD BACTEC™ MGIT™ 960 system, and stool sample were tested with Xpert-Ultra assay and smear microscopy. Demographic data, clinical information and laboratory test results were electronically recorded in password-protected TB programmatic and laboratory databases separately, and then data was combined to perform analysis.

### Sample collection

Trained healthcare workers provided verbal instructions to participants or their caregivers on proper collection of sputum and stool samples using a step-by-step workflow in accordance with the Standard Operating Procedures. Two sputum samples including spot and early morning sputum samples of at least 3 ml or one NGA sample with minimum 1 mL in case of sputum cannot be expectorated were collected.

Participants themselves or caregivers were instructed to take it from first bowel movement of the day either at home or study sites and reminded to avoid contamination with urine or disinfectants. Pre-labelled sputum containers, clear sealable plastic bags, clean wooden sticks were provided to use in sample collection. Participants were instructed to collect up to halfway of stool container to be able to obtain sufficient amount of stool with approximately 3-5 grams for retesting in case of unsuccessful first test.

### Sample storage and transport

Samples were placed in closed containers within clean plastic bags and transported at ambient temperature to the study sites on the same day of collection. They were stored at 2–8 °C, protected from direct sunlight, and transported to the SMRU main laboratory daily.

### Sample processing and procedures

#### Microscopy

The sputum and stool samples were tested following the SMRU’s standard operating procedures. Briefly, after reviewing reception/rejection criteria and registered samples to the laboratory. Samples were stored at 2-8°C until testing. Samples were checked for macroscopic appearances such as blood-stained, muco-purulent, purulent (with saliva), or saliva. Smears were then prepared and stained using Ziehl Neelsen method. Microscopic examination was performed using 100x objective lens. Smear results, including grading, were reported [15].

#### Xpert MTB/RIF Ultra Assay

Stool types were identified using Bristol stool chart, and Simple One-Step stool processing method, recommended by WHO was applied for sample processing. For solid stool, 0.8 g stool was transferred into sample reagent bottle, shaken for 30 seconds and stood for 10 minutes. This step was repeated again, then 2 mL of debris-free supernatant was transferred into the Xpert-Ultra cartridge for testing according to the manufacturer’s instructions. For liquid stool, 2 mL of sample reagent was removed and disposed. Then 2 mL of liquid stool was then transferred into the sample reagent bottle before performing the similar procedure as described in solid stool.

For sputum samples, 1 mL of early morning sample was transferred into the labeled bijou and mixed with 2 mL of sample reagent. It was then inverted for 10 seconds and stood for 10 minutes. The tube was further inverted for next 10 seconds and left stood for another 5 minutes before loading 2 mL of sample/sample reagent mix into the Xpert-Ultra cartridge. The cartridge was then loaded onto the machine and processed following the manufacturer’s instructions. Semi-quantitative results for both samples were reported and recorded in the SMRU microbiology database [16]. Both smear microscopy and Xpert-Ultra assay for sputum and stool specimen were performed at the SMRU main laboratory, Mae Ramat district, Tak province, Thailand. Sputum smear microscopy and Xpert-Ultra assay result were not blinded to performers of the stool smear and Xpert-Ultra assay.

#### MTB culture

Early morning sputum samples (≥1 mL) were sent to the International Organization for Migrant laboratory, Mae Sot district, Tak province, Thailand for MTB culture. All selected samples are sealed in plastic bags and delivered with clearly labeled cool box, keeping temperature between 2-8 °C. After decontamination with NaOH/N-acetyl L-cysteine, sample was incubated in MGIT culture media (BACTEC MGIT 960 System; Becton Dickinson, Franklin Lakes, NJ, USA). Negative growth was confirmed after 8 weeks of incubation. First and second line anti-TB drug sensitivity tests were also proceeded for positive samples. The laboratory staff were not informed about clinical information and results of other diagnostic methods.

#### Sample size

A minimal sample size of 120 participants was required using the Buderer’s formula, assuming a sensitivity of 90% and specificity of 96% for Xpert-Ultra assay using stool samples, at a 10% level of precision [2, 17]. TB prevalence was predicted to be 30% among the study population considering 10% incomplete data. Participants under 18 years were excluded due to different nature of disease development and potential lower bacillary load in children compared to adults, giving total number of 113 adult participants in final analysis.

#### Statistical analysis

Data were entered in Microsoft Access 2016 and analyzed using IBM SPSS Statistics, version 29.0 (IBM Corp., Armonk, NY, USA). Among total enrolled 120 participants, individuals aged less than 18 years were excluded in final analysis for two reasons – the difference in nature of TB disease development between children and adults, pauci-bacillary versus multi-bacillary disease and unexpectedly low number of children and adolescent participants (7/120).

Demographic and clinical characteristics of the study participants were summarized using frequencies and percentages for categorical variables and compared using a chi-squared test, whereas continuous variable like age that approximately normally distributed in both groups was presented by means with standard deviation and compared by Student’s independent-samples t-test. Statistical significance was set as p-value less than 0.05.

Based on sputum MTB culture as gold standard, diagnostic performance measures of index tests (Xpert-Ultra assay and smear microscopy) for stool and sputum samples (sensitivity, specificity, positive, and negative predictive values and their 95% confidence intervals) were calculated with 2x2 contingency table using the MedCalc diagnostic test evaluation calculator (MedCalc Software Ltd., Belgium). In addition, TB case detection rates with different diagnostic methods were presented, and statistical differences between the two types of specimens (stool versus sputum) was determined using McNemar’s test.

### Ethical considerations

The study was approved from the Oxford Tropical Research Ethics Committee (Ref: 570-22) and local Community Ethics Advisory Board of Tak province (Ref: CEAB-2022-016). Written informed consent was obtained from adult participants or parents or legal guardians of children.

## Results

### Demographic and clinical characteristics of study participants

In total 163 participants were screened for enrolment, of which 4 participants were ineligible because they had no presumptive TB symptoms. Among remaining 159 participants, 39 participants were excluded in final analysis due to the following reasons: no sputum culture results in 10 participants, no sputum Xpert-Ultra assay results in 9 participants, no spot sputum smear microscopy result in one participant, no stool specimens in 16 participants and missing case record forms in 3 participants. In addition, 7 participants under 18 years of age were also excluded due to potential difference in bacillary load and nature of disease and low number of childhood and adolescent study participants. Therefore, a total of 113 individuals with presumptive TB were included for the final analysis.

Among these participants, 63.7 % (72/113) were male, and mean age was 44.7 ± 16.4 years. Majority of the study participants (76.1 %) were migrants, and half of them were Burmese ethnic (48.7%). While over 70 % of the participants came from the outpatient department of respiratory diseases, the remaining participants were hospitalized.

Analyzing the presenting symptoms, cough was the most commonly reported symptoms with 92.9%, followed by weight loss with 65.5%, fever with 51.3% and night sweat about 39.6%. While 14 participants (12.6%) had history of contact with TB patients before, 18 participants (16.1%) reported that they had previous history of TB treatment. Regarding risk factors for TB, 36 participants (32.4%) and 50 participants (44.6%) reported alcohol drinking and smoking respectively. In comorbidities, 14.4% and 8.0 % of the participants were found out to have diabetes and HIV.

There were 33 bacteriologically confirmed TB (BC-TB) patients diagnosed by sputum smear microscopy and/or Xpert-Ultra assay and/or culture on sputum samples. The mean age of these patients was 43.1 ± 17.2 and 26 (78.8%) patients were males. Among them, 2 (6.3 %) patients were HIV-positive, 11 (34.4%) patients had HIV negative status, and 19 (59.4%) had an unknown HIV status (Table 1).

**Table 1.** Demographic and clinical characteristics of the study population. BC-TB was defined as positive result on sputum specimen of smear microscopy and/or Xpert-Ultra assay and/or MTB culture.

| Characteristics | Total participants (n=113) | Participants with BC-TB (n= 33) | Participants without BC-TB (n= 80) | P-value |
| --- | --- | --- | --- | --- |
| Age, mean ± SD | | 43.1 ± 17.2 | 45.4 ± 16.1 | 0.507 |
| **Sex** |  |  |  |  |
| Male | 72 (63.7%) | 26 (78.8%) | 46 (57.5%) | **0.032** |
| Female | 41 (36.3%) | 7 (21.2%) | 34 (42.5%) |  |
| **Migrant** |  |  |  |  |
| Yes | 86 (76.1%) | 22 (66.7%) | 64 (80.0%) | 0.131 |
| No | 27 (23.9%) | 11 (33.3%) | 16 (20.0%) |  |
| **History of TB contact** |  |  |  |  |
| Yes | 14 (12.6%) | 5 (15.2%) | 9 (11.5%) | 0.600 |
| No | 97 (87.4%) | 28 (84.8%) | 69 (88.5%) |  |
| **Cough** |  |  |  |  |
| Yes | 105 (92.9%) | 32 (97.0%) | 73 (91.3%) | 0.281 |
| No | 8 (7.1%) | 1 (3.0%) | 7 (8.8%) |  |
| **Weight loss** |  |  |  |  |
| Yes | 72 (65.5%) | 26 (81.3%) | 46 (59.0%) | **0.026** |
| No | 38 (34.5%) | 6 (18.8%) | 32 (41.0%) |  |
| **Fever** |  |  |  |  |
| Yes | 58 (51.3%) | 21 (63.6%) | 37 (46.3%) | 0.093 |
| No | 55 (48.7%) | 12 (36.4%) | 43 (53.8%) |  |
| **Night Sweat** |  |  |  |  |
| Yes | 38 (39.6%) | 13 (44.8%) | 25 (37.3%) | 0.489 |
| No | 58 (60.4%) | 16 (55.2%) | 42 (62.7%) |  |
| **Smoking** |  |  |  |  |
| Yes | 50 (44.6%) | 19 (59.4%) | 31 (38.8%) | **0.047** |
| No | 62 (55.4%) | 13 (40.6%) | 49 (61.3%) |  |
| **Alcohol** |  |  |  |  |
| Yes | 36 (32.4%) | 16 (51.6%) | 20 (25.0%) | **0.007** |
| No | 75 (67.6%) | 15 (48.4%) | 60 (75.0%) |  |
| **Diabetes** |  |  |  |  |
| Yes | 16 (14.4%) | 3 (9.4%) | 13 (16.5%) | 0.500 |
| No | 76 (68.5%) | 22 (68.8%) | 54 (68.4%) |  |
| Unknown | 19 (17.1%) | 7 (21.9%) | 12 (15.2%) |  |
| **HIV** |  |  |  |  |
| Yes | 9 (8.0%) | 2 (6.3%) | 7 (8.8%) | 0.896 |
| No | 39 (34.8%) | 11 (34.4%) | 28 (35.0%) |  |
| Unknown | 64 (57.1%) | 19 (59.4%) | 45 (56.3%) |  |
| **Past History of TB** |  |  |  |  |
| Yes | 18 (16.1%) | 2 (6.1%) | 16 (20.3%) | 0.062 |
| No | 94 (83.9%) | 31 (93.9%) | 63 (79.7%) |  |

**Figure 1.**
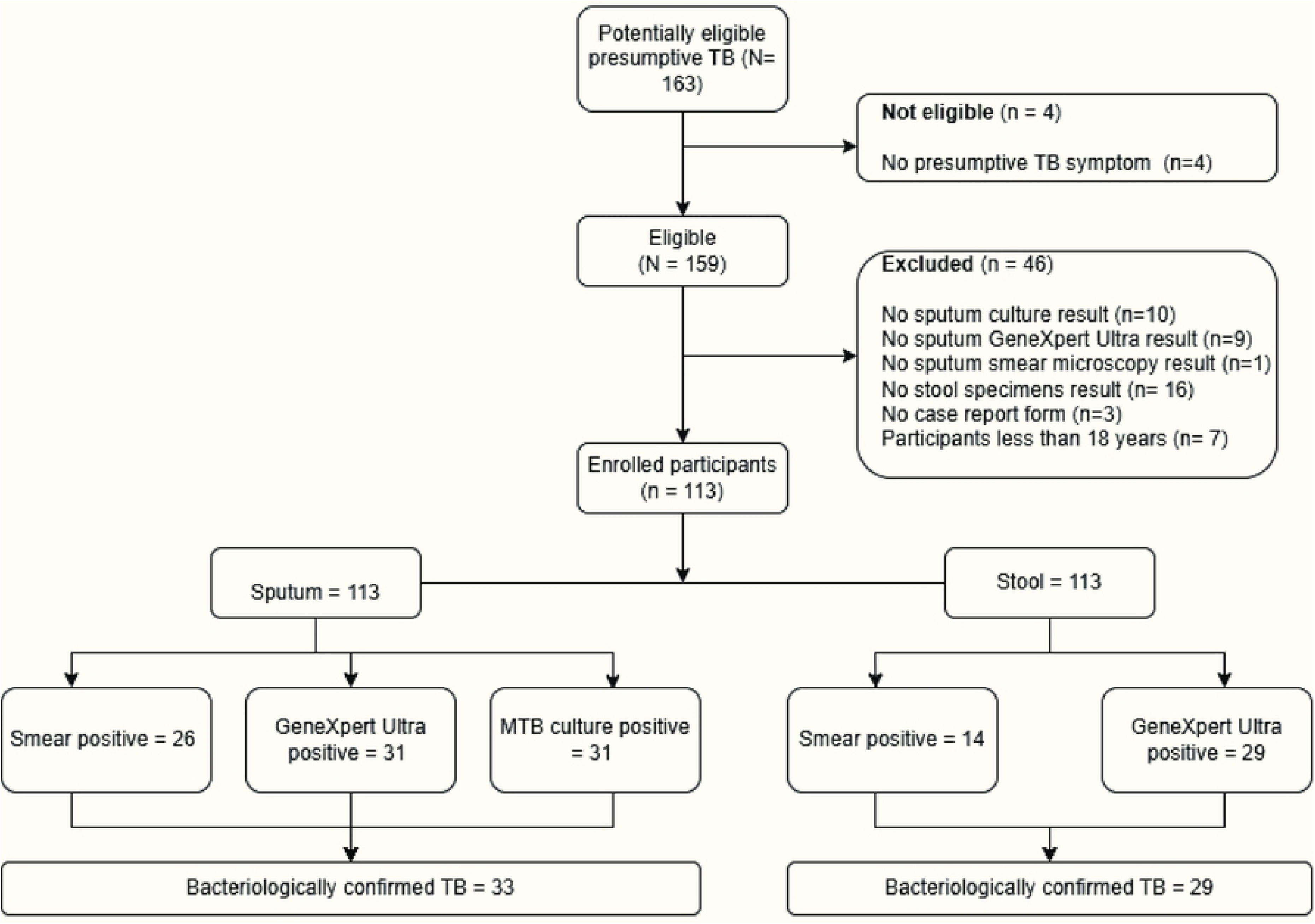
Diagnosis of TB in sputum and stool specimens. Statistical differences of demographic and clinical characteristics among participant groups with BC-TB and those without are described in the **Table 1**. In the group without BC-TB, 7 participants with clinically diagnosed TB were included.

### *Mycobacterium tuberculosis* complex detection rate by different TB diagnostic methods

The MTB detection rate by different diagnostic methods were presented in **Table 2**. On the MTB culture in sputum, the MTB complex detection rate was consistent to the sputum Xpert-Ultra test, 27.43% (31/113). There were two discordant results between these two tests. While non-tuberculosis mycobacteria (NTM) growth was identified among 11 out of 113 (9.7 %) sputum, no growth was discovered in 71 out of 113 (62.8%) sputum samples. These two groups (72.6 %) were collectively categorized as negative for MTB on culture.

**Table 2:** Mycobacterium tuberculosis complex detection rate by different TB diagnostic methods.

| DIAGNOSTIC METHOD | POSITIVE (% , NO) | NEGATIVE (% , NO) |
| --- | --- | --- |
| ***SPUTUM CULTURE*** | 27.4% (31) | 72.6% (82 – included 11 NTM) |
| ***SPUTUM XPERT-ULTRA*** | 27.4% (31) | 72.6% (82) |
| ***STOOL XPERT-ULTRA*** | 25.7% (29) | 74.3% (84) |
| ***SPUTUM SMEAR MICROSCOPY*** | 23.0% (26) | 77.0% (87) |
| ***STOOL SMEAR MICROSCOPY*** | 12.4% (14) | 87.6% (99) |

The MTB DNA detection rate by the Xpert-Ultra assay was also 27.4 % (31/113) in sputum samples and 25.7 % (29/113) in stool samples respectively. All positive results on stool samples were concordant with positive results on the sputum culture and Xpert-Ultra tests. Although TB case detection rates on stool specimen was 1.77 % (95% CI, −4.20 % to 0.66 %) lower than that of sputum specimen, these were not statistically different (McNemar test, p = 0.5). Three rifampicin resistant cases were detected in both sputum and stool samples. In sputum samples, two out of 31 (6.4%) Xpert-Ultra positive TB cases were found as very-low level bacterial load. No case with trace result was included in this analysis although one case was found and excluded due to indeterminate result of rifampicin resistant detection. The remaining 93.6% (29/31) were appeared with stronger gradings of high, medium and low levels. In stool samples, five out of 29 (17.2%) positive cases were shown as with very-low level of grading while the remaining 24 cases (82.8%) were positive with medium and low level. There was also no trace-level positive case.

By sputum smear microscopy, overall acid-fast bacilli (AFB) detection rate was 23.0% (26/113) with 22.1% (25/113) from early morning samples and 20.4% (23/113) from spot samples. When case detection rate using sputum smear microscopy was compared to the Xpert-Ultra assay in stool specimens, there was difference in -2.65 % (95% CI, −0.31 % to 5.62 %), showing no statistically significant difference between the tests (McNemar test, p=0.15). The detection rate was lowest by stool smear microscopy test with only 12.4% (14/113) and no discordant result was discovered compared to other tests.

### Performance of Xpert MTB/RIF Ultra Assay on stool specimen

With gold standard method of MTB culture on sputum specimen, diagnostic accuracy of Xpert-Ultra assay on stool and sputum specimens for detection of TB was compared in **Table 3**. The sensitivity of stool and sputum sample was 90.32% (95% CI, 74.25% to 97.96%) and 93.55% (95% CI, 78.58% to 99.21%) and specificity was 98.78% (95% CI, 93.39% to 99.97%) and 97.56% (95% CI, 91.47% to 99.70%) respectively. Despite lower absolute difference in sensitivity of Xpert-Ultra assay in stool (difference, -3.23 %; 95% CI: -9.46 % to 3.00 %), there was no statistically significant difference between the two sample types (McNemar test, p=1.0). Absolute difference in specificity of the Xpert-Ultra assay in stool specimens was 1.22 % (95% CI: -1.12 % to 3.59 %), showing no evidence of statistically difference (McNemar test, p=1.0). When diagnostic performance of the Xpert-Ultra assay in stool was compared to sputum smear microscopy method, there was no statistically different (McNemar test, p=0.25 for sensitivity and specificity, p=1.0).

**Table 3.**
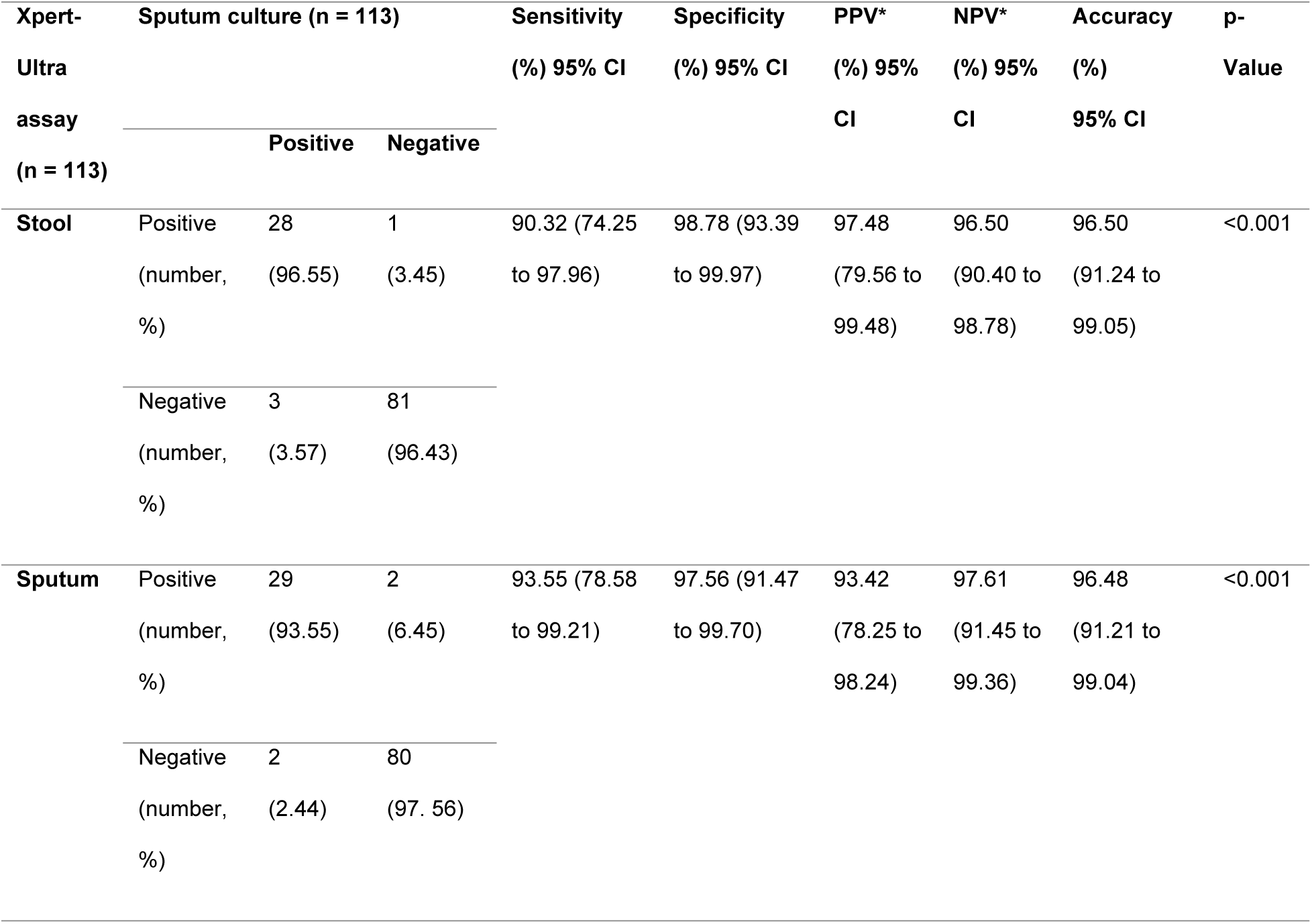
Performance of Xpert-Ultra assay for identification of MTB in stool and sputum specimens using sputum MTB culture as the gold standard. (*) disease prevalence was calculated based on the disease prevalence estimated from the current study findings.

### Performance of smear microscopy on stool specimen

The sensitivity of the stool microscopy method compared to the gold standard method was 41.94% (95% CI, 24.55% to 60.92%), which was 38.71 % (95% CI, -55.85 % to -21.57 %) lower than that of the conventional sputum microscopy method in which it had 80.65% (95% CI, 62.53% to 92.55%). Statistical difference between two methods was observed (McNemar test, p < 0.001). Specificity was consistent with sputum smear microscopy with 98.78% (95% CI, 93.39% to 99.97%) (**Table 4**).

**Table 4.**
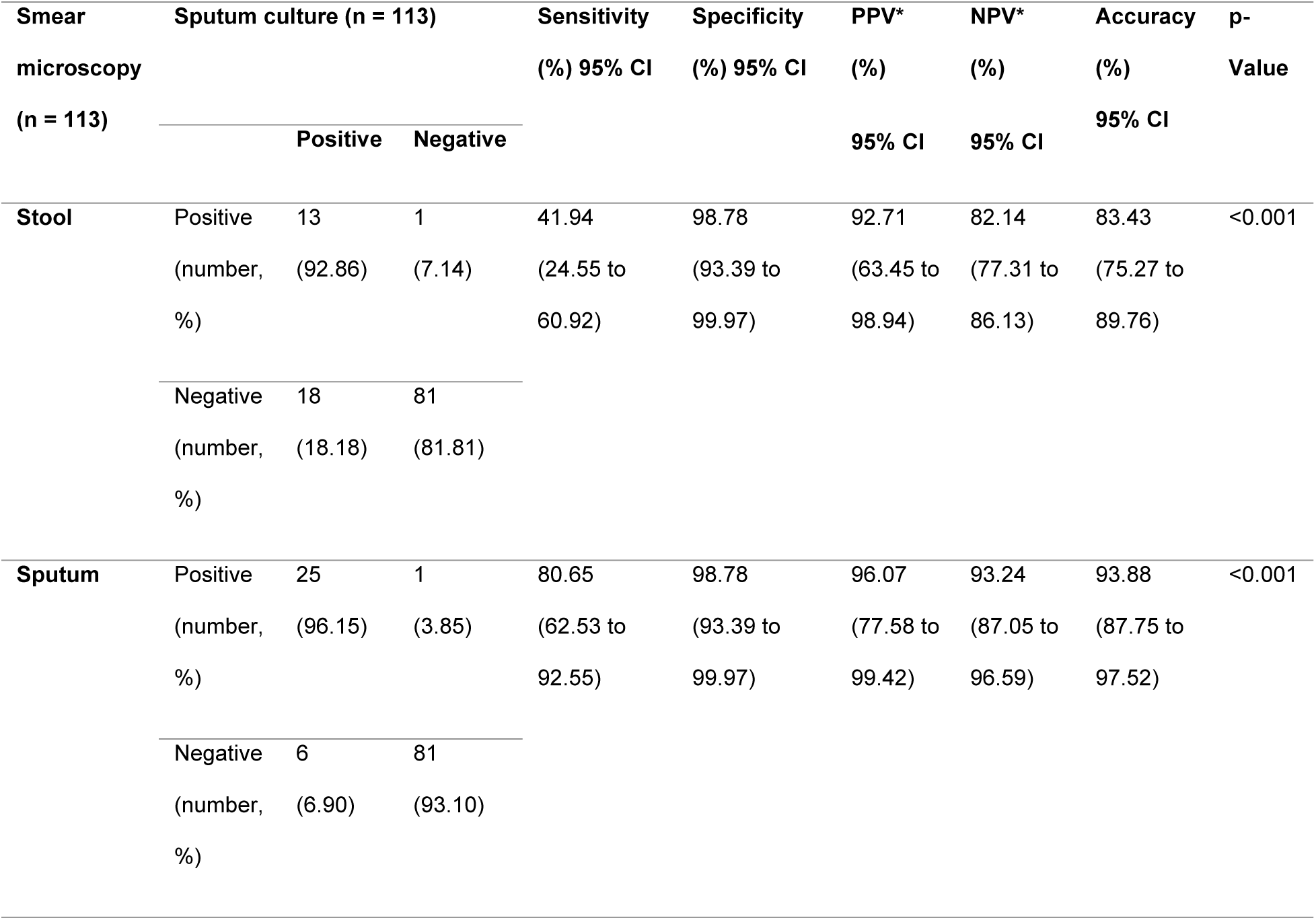
Performance of smear microscopy for identification of AFB in stool and sputum specimens using sputum MTB culture as the gold standard. (*) disease prevalence was calculated based on the disease prevalence estimated from the current study findings.

## Discussion

To the best of our knowledge, our study is the first study to evaluate performance of the Xpert-Ultra assay with stool specimens among adult presumptive TB patients living in border area with high TB burden and generalized HIV epidemic situation in Thailand [18].

With sputum MTB culture as gold standard, the performance of stool Xpert-Ultra test was higher than Yu et al. study in China that reported sensitivity and specificity of 78.43% and 77.22% respectively [19]. In that study, PLHIV, in whom disseminated and extra-pulmonary TB such as gastro-intestinal (GI) TB are common, was not included; it may contribute to lower sensitivity in their study. The sensitivity in our study was consistent to findings of previous studies conducted in Uganda with high HIV prevalence, and in Bangladesh where HIV prevalence was low [20, 21]. However, the lower specificity (91%) in the Uganda study can probably be due to cross-reaction with NTM while maintaining high bacterial load by processing collected stool sample with a sample transport reagent used for sputum decontamination. The high sensitivity of stool Xpert-Ultra test in our study may also be due to having greater bacilli load in the study participants with BC-TB patients as shown by sputum Xpert-Ultra MTB-DNA load without any trace result and very small proportion with very-low level of qualitative grading.

The findings of the highest performance by the Xpert-Ultra assay on MTB detection in sputum samples was comparable to a study findings in China (sensitivity, 92.26% and specificity, 96.30%) [22], indicating highly beneficial method for diagnosis of TB among presumptive TB patients with productive cough. Sensitivity of our study was higher than Dorman S. E. et al., study [23] because TB case in that study was defined as positive sputum culture from at least one of four sputum cultures from each participant, thereby enhancing accuracy of the gold standard test for identification of true positives and subsequently having lower sensitivity in index test. In Uganda, Abdulwahab Sessolo reported comparable sensitivity of 95% and lower specificity of 90% compared to our study findings [20]. The lower specificity in the Uganda could be explained by higher HIV proportion in their study than in our study, in whom extra-pulmonary TB is common, and this limitation subsequently might be a reason of obtaining lower specificity of index test with high number of false positive cases. Based on these findings, the Xpert-Ultra assay on sputum specimen showed the highest diagnostic performance compared to alternative diagnostic methods in our study.

With smear microscopy examination, performance of our study on stool specimen is consistent to the study finding reported by Abdulwahab Sessolo [20], in which a sample transport reagent is used for sputum decontamination. Our study finding on stool specimen had higher sensitivity than that of a study finding in China [19], with 11.94 % of sensitivity. It is probably due to higher proportion of PLHIV in our study, and thus possibility of having extrapulmonary TB in GI tract and excreting higher bacilli load in stool specimen which could lead to higher sensitivity by smear microcopy. However, as smear microscopy cannot differentiate MTB and NTM, it is noted smear microscopy can produce false positive result [24]. In sputum smear method, our study found similar performance reported by Abdulwahab Sessolo and Rahman [20, 21]. A higher sensitivity was achieved compared to study findings reported by Sun Lin [25] and Liu Xu-hui [26] (25% and 27% respectively). This difference can be explained by the fact that children were included in those studies and were noted as population with low bacterial load in collected specimen.

In our study, although the AFB were detected in the stool, positive case detection rate by the smear microscopy in stool specimen was lower than in sputum with 12.39% and 23.01% respectively. This finding was similar to the findings reported by Odilon D. Kabore [27]. Moreover, all stool AFB positive results were concordant with the stool Xpert-Ultra positive results. Similarly in the study findings in China and Uganda [19, 20], stool Xpert-Ultra assay had more case detection rate than smear microscopy on stool specimen, supporting our study findings. With these evidences and significant statistical difference in sensitivity of smear microscopy method between stool and sputum specimens, smear microscopy on stool specimen had no added value to be deployed as parallel test for TB diagnosis.

Recently WHO recommended tongue swab as an alternative sample for PTB diagnosis when respiratory samples are not available. The meta-analysis on tongue swab tested by PCR assays conducted by the WHO guideline development groups reported the pool sensitivity of 71.4% (95% CI, 67.4% to 75.1%) and specificity of 98.8% (95% CI, 97.6% to 99.4%) among adolescents and adults with presumptive pulmonary TB [28]. The systematic review on stool sample use for PTB diagnosis with various PCR assays exhibited varied sensitivity within the range of 69.7% to 100% and specificities from 69.8% to 100% [11], indicating that stool sample test has the potential to use as alternative sample like tongue swab if specificity is improved. Moreover, the WHO recommended minimum 80% of sensitivity and 98% of specificity as target product profiles for diagnostic performance in developing low-complexity assays in non-sputum sample [29]. In our study, the Xpert-Ultra assay in stool was achieved the benchmarked level of diagnostic performance.

In conclusion, the Xpert-Ultra performance in stool specimen was similar to that of sputum with overall accuracy of 96.5% (95% CI, 91.2% to 99.1%) in stool and 96.5% (95% CI, 91.2% to 99.0%) in sputum respectively. In addition, all Xpert-Ultra stool sample positive results were concordant with positive sputum sample in our study. These findings support other studies that Xpert-Ultra stool can be considered as an alternative method for diagnosis of TB among adults who are hard to expectorate sputum and where microbiological laboratories for culture are not available. Though culture and Xpert-Ultra tests can equally detectable to TB, the prolong turn-around time of culture result is not pragmatic to apply as an initial diagnostic tool and could delay treatment initiation.

## Limitations

There are some limitations in our study. Firstly, as pediatric population who can be potentially beneficial from this study were excluded, as it might limit generalizability of study findings in the specific sub-population. Secondly, we did not confirm for the possibility of gastro intestinal TB in a participant whose smear microscopy and Xpert-Ultra assay results on both sputum and stool samples were found as positive while NTM was detected on corresponding sputum culture. Thirdly, we could not understand applicability of the Xpert-Ultra assay on stool specimen for detection of Rifampicin resistance because we identified only three rifampicin resistant cases with Xpert-Ultra assay on stool and sputum samples. Finally, we found that there was no significant statistical difference in performance of Xpert-Ultra assay between stool and sputum; however, larger samples are required to confirm equivalence and to inform more accurate estimates of diagnostic performance.

## Conclusion

Although WHO recommends concurrent testing using Xpert-Ultra assay on stool and respiratory samples in children with presumptive TB symptoms, bacteriological diagnosis of PTB among adults mainly rely on sputum sample examination – the test result varies with quality and volume of sputum sample. In our study, among adult presumptive TB participants, stool sample using Xpert-Ultra assay showed comparable diagnostic performances with sputum sample. Due to slightly lower diagnostic accuracy of the Xpert-Ultra assay on stool sample and no data could be generated for PTB patients with trace bacilli load, we cannot suggest to replace Xpert-Ultra assay and MTB culture on sputum specimen; however, we suggest the fact that it can has a potential clinical role as an alternative method in diagnosis of PTB in adults when sputum specimen is difficult to obtain. Additionally, our study added more data about diagnostic performances on stool sample using Xpert-Ultra assay and microscopy, compared to routine diagnostic sample among vulnerable migrants living in generalized HIV epidemic area.

## Data Availability

The data supporting the findings of this study are available in accordance with the data sharing policy of the Mahidol Oxford Tropical Medicine Research Unit (MORU). Researchers may request access by contacting the MORU Data Access Committee. Requests will be evaluated based on scientific merit and ethical considerations.

## Acknowledgements

We would like to acknowledge the Welcome Trust for providing laboratory support for testing the Xpert-Ultra assay and culture. In addition, we thank to healthcare workers and laboratory staff from the SMRU and MTC for providing required supports in study recruitment. The authors gratefully acknowledge to Dr. Lei Swe, former research clinician, SMRU and Mr. James Reed, visiting student from Stanford University, USA, for their contributions to the conceptualization and laboratory testing in this study. Finally, we deeply acknowledge the participants provided consent to involve in the study.

## Citation

**Htet Ko Ko A, Sein ST, Wanitda W, Aung PP, Francois N. (2026) Diagnostic Performance of Xpert MTB/RIF Ultra Assay for Tuberculosis in Stool specimens among Adult presumptive TB patients in a Generalized HIV Epidemic Setting.**

## Author Contributions

Conceptualization: Htet Ko Ko A, Sein ST, Wanitda W, Aung PP, Francois N.

Data curation: Htet Ko Ko A.

Formal Analysis: Htet Ko Ko A, Sein ST.

Funding acquisition: Francois N.

Investigation: Wanitda W.

Methodology: Htet Ko Ko A, Wanitda W, Aung PP, Francois N.

Project administration: Htet Ko Ko A.

Supervision: Francois N.

Writing – Original Draft Preparation: Htet Ko Ko A.

Writing – Review & Editing: Htet Ko Ko A, Sein ST, Wanitda W, Aung PP, Francois N.

## References

1. Global Tuberculosis Report 2024. Geneva: World Health Organization; 2024. Licence: CC BY-NC-SA 3.0 IGO.

2. WHO consolidated guidelines on tuberculosis. Module 3: diagnosis - rapid diagnostics for tuberculosis detection, 2021 update. Geneva: World Health Organization; 2021.Licence: CC BY-NC-SA 3.0 IGO.

3. Giang DC, Duong TN, Ha DTM, Nhan HT, Wolbers M, Nhu NTQ, et al. Prospective evaluation of GeneXpert for the diagnosis of HIV-negative pediatric TB cases. BMC infectious diseases. 2015;15(1):70.

4. Esmail A, Pooran A, Sabur NF, Fadul M, Brar MS, Oelofse S, et al. An optimal diagnostic strategy for tuberculosis in hospitalized HIV-infected patients using GeneXpert MTB/RIF and Alere determine TB LAM Ag. Journal of clinical microbiology. 2020;58(10):10.1128/jcm.01032-20.

5. Sharma MV, Arora VK, Anupama N. Challenges in diagnosis and treatment of tuberculosis in elderly. Indian Journal of Tuberculosis. 2022;69:S205–S8.

6. Caraux-Paz P, Diamantis S, de Wazières B, Gallien S. Tuberculosis in the elderly. Journal of Clinical Medicine. 2021;10(24):5888.

7. Cox H, Workman L, Bateman L, Franckling-Smith Z, Prins M, Luiz J, et al. Oral Swab Specimens Tested With Xpert MTB/RIF Ultra Assay for Diagnosis of Pulmonary Tuberculosis in Children: A Diagnostic Accuracy Study. Clin Infect Dis. 2022;75(12):2145–52.

8. Fauci AS, Eisinger RW. Reimagining the Research Approach to Tuberculosis(†). Am J Trop Med Hyg. 2018;98(3):650–2.

9. Hanrahan CF, Dansey H, Mutunga L, France H, Omar SV, Ismail N, et al. Diagnostic strategies for childhood tuberculosis in the context of primary care in a high burden setting: the value of alternative sampling methods. Paediatrics and international child health. 2019;39(2):88–94.

10. WHO consolidated guidelines on tuberculosis. Module 3: diagnosis. Geneva: World Health Organization; 2025. Licence: CC BY-NC-SA 3.0 IGO.

11. Sultana S, Afrin S, Hasan M, Ansar A, Saif-Ur-Rahman K. Stool specimen for diagnosis of pulmonary tuberculosis in adults: a systematic review. BMJ open. 2023;13(4):e062135.

12. Hemhongsa P, Tasaneeyapan T, Swaddiwudhipong W, Danyuttapolchai J, Pisuttakoon K, Rienthong S, et al. TB, HIV-associated TB and multidrug-resistant TB on Thailand’s border with Myanmar, 2006-2007. Trop Med Int Health. 2008;13(10):1288–96.

13. Mae Tao Clinic. Annual report. Mae Sot: Mae Tao Clinic; 2024 [Available from: https://maetaoclinic.org/publications/annual-reports/.

14. WHO consolidated guidelines on tuberculosis. Module 2: screening-systematic screening for tuberculosis disease. Geneva: World Health Organization; 2021. Licence: CC BY-NC-SA 3.0 IGO.

15. Global Laboratory Initiative. Laboratory diagnosis of tuberculosis by sputum microscopy. Adelaide: SA Pathology; 2013. 84. p.

16. Cepheid. Xpert MTB-RIF Ultra English Package Insert, Rev. D. Sunnyvale (CA): Cepheid; 2023.

17. Buderer NMF. Statistical methodology: I. Incorporating the prevalence of disease into the sample size calculation for sensitivity and specificity. Academic Emergency Medicine. 1996;3(9):895–900.

18. Press B. Migrants’ Health and Vulnerability to HIV/AIDS in Thailand: PHAMIT/Raks Thai Foundation; 2009 [Available from: https://www.aidsdatahub.org/sites/default/files/resource/migrant-health-and-hiv-vulnerability-thailand.pdf.

19. Yu X, Wang F, Ren R, Dong L, Xue Y, Zhao L, et al. Xpert MTB/RIF Ultra Assay Using Stool: an Effective Solution for Bacilli Identification from Adult Pulmonary Tuberculosis Suspects without Expectorated Sputum. Microbiology Spectrum. 2023;11(4):e01265–23.

20. Sessolo A, Musisi E, Kaswabuli S, Zawedde J, Byanyima P, Sabiiti W, et al. Diagnostic accuracy of Xpert MTB/RIF Ultra and culture assays to detect Mycobacterium Tuberculosis using OMNIgene-sputum processed stool among adult TB presumptive patients in Uganda. Plos one. 2023;18(4):e0284041.

21. Rahman SM, Maliha UT, Ahmed S, Kabir S, Khatun R, Shah JA, et al. Evaluation of Xpert MTB/RIF assay for detection of Mycobacterium tuberculosis in stool samples of adults with pulmonary tuberculosis. PLos one. 2018;13(9):e0203063.

22. Wang G, Huang M, Jing H, Jia J, Dong L, Zhao L, et al. The practical value of Xpert MTB/RIF Ultra for diagnosis of pulmonary tuberculosis in a high tuberculosis burden setting: a prospective multicenter diagnostic accuracy study. Microbiology Spectrum. 2022;10(4):e00949–22.

23. Dorman SE, Schumacher SG, Alland D, Nabeta P, Armstrong DT, King B, et al. Xpert MTB/RIF Ultra for detection of Mycobacterium tuberculosis and rifampicin resistance: a prospective multicentre diagnostic accuracy study. The Lancet infectious diseases. 2018;18(1):76–84.

24. Levy H, Feldman C, Sacho H, van der Meulen H, Kallenbach J, Koornhof H. A reevaluation of sputum microscopy and culture in the diagnosis of pulmonary tuberculosis. Chest. 1989;95(6):1193–7.

25. Sun L, Liu Y, Fang M, Chen Y, Zhu Y, Xia C, et al. Use of Xpert MTB/RIF Ultra assay on stool and gastric aspirate samples to diagnose pulmonary tuberculosis in children in a high-tuberculosis-burden but resource-limited area of China. International Journal of Infectious Diseases. 2022;114:236–43.

26. Liu X-h, Xia L, Song B, Wang H, Qian X-q, Wei J-h, et al. Stool-based Xpert MTB/RIF Ultra assay as a tool for detecting pulmonary tuberculosis in children with abnormal chest imaging: a prospective cohort study. Journal of Infection. 2021;82(1):84–9.

27. Kaboré OD, Millogo A, Sanogo B, Birba E, Poda A, Nacro B, et al. Analytical performances of the Xpert MTB/RIF assay using stool specimens to improve the diagnosis of pulmonary tuberculosis in Burkina Faso, a tuberculosis endemic country. PLoS One. 2023;18(7):e0288671.

28. Organization WH. Near point-of-care tests, tongue swabs and sputum pooling for tuberculosis diagnosis 2026 [Available from: https://www.who.int/teams/global-programme-on-tuberculosis-and-lung-health/diagnosis-treatment/npoc-tongue-swabs-and-sputum-pooling-for-tb.

29. Organization WH. Target product profiles for tuberculosis diagnosis and detection of drug resistance. 2024.

